# The early impact of vaccination against SARS-CoV-2 in Region Stockholm, Sweden

**DOI:** 10.1101/2021.09.02.21263046

**Authors:** Catherine Isitt, Daniel Sjöholm, Maria-Pia Hergens, Fredrik Granath, Pontus Nauclér

**Affiliations:** Department of Infectious Diseases, Karolinska University Hospital, Stockholm, Sweden; Department of Medicine Solna, Division of Infectious Diseases, Karolinska Institutet, Sweden; Department of Medicine Solna, Clinical Epidemiology Unit, Karolinska Institutet, Karolinska University Hospital, Sweden; Department of Communicable Disease Control and Prevention, Stockholm County Council, Stockholm, Sweden

**Keywords:** COVID-19, vaccines, SARS-CoV2, cohort study, Long-term-care-facilities

## Abstract

Vaccination against SARS-CoV-2 started in Region Stockholm, Sweden in December 2020 with those in long-term care facilities or receiving home care vaccinated first followed by those aged over 80 years. In this population-based, retrospective cohort study, we performed a Poisson regression to model the expected incidence of infections and deaths which we compared to the observed incidence and compared this to an unvaccinated control group of those aged 18-79 years. The aim of this study was to measure the early impact of the vaccination programme in Region Stockholm.

Infections and deaths reduced substantially amongst the first two groups targeted for SARS-CoV-2 vaccination with an estimated total 3112 infections prevented, and 854 deaths prevented in these two groups from 4 weeks after the introduction of vaccination through to 2^nd^ May 2021.

## Introduction

The Swedish COVID-19 vaccination programme prioritises those most at risk of severe disease from SARS-CoV-2 infection first with the aim of protecting those most vulnerable and safeguarding the healthcare system. The programme is divided into four phases: 1) adults living in long-term care facilities (LTCF), adults receiving home care and their household contacts, and healthcare staff working with this population, 2) adults over the age of 65 years as well as adults of any age who receive dialysis or are transplant recipients, with the oldest invited for vaccination first, 3) adults with other risk factors, 4) adults without risk factors over 18 years of age[1]. In Stockholm, vaccination of Phase 1 began on 27^th^ December and vaccination of Phase 2 started on 8^th^ March 2021 with adults ≥80 years invited for vaccination first[2]. Adults ≥75 years were invited starting on 29^th^ March 2021[2, 3].

To assess the early impact of the vaccination programme in Stockholm, we performed a population-based, retrospective cohort study to investigate the incidence of PCR-confirmed SARS-CoV-2 as well as deaths within 30 days of a PCR-confirmed SARS-CoV-2 diagnosis, in the cohorts targeted for vaccination compared to a control group not targeted for vaccination.

## Methods

### Study population

The Stockholm COVID-19 cohort study collects data from all residents living in Region Stockholm from 2015 onwards. We extracted data from the VAL database which collates information from more than ten other healthcare related databases in Region Stockholm and which has been previously described[4]. Age, and coding for resident in LTCF and home care were used to define the two groups first targeted for vaccination, those in LTCF/home care and those aged ≥80 years, plus a third group, those aged 18-79 years who were used as a composite unvaccinated control. Linkage to SmiNet, the Public Health Agency of Sweden notifiable infection reporting tool, was performed using unique personal identification numbers that each resident in Sweden has, to extract data on SARS-CoV-2 PCR-positive cases.

The study was approved by the Swedish Ethical Review Authority.

### Statistics

Analyses were restricted to 31^st^ August 2020 to 2^nd^ May 2021 in order to encompass the second and third waves of COVID-19 in Stockholm. Data from the first wave were not used given the lack of widely available testing at the start of the pandemic. Three mutually exclusive groups were defined, those in LTCF or receiving home care, those aged 80+ years and a composite control of those aged 18-79 years. The time point set for post-vaccination was 4 weeks after the start of vaccination, to account for the fact that the protective effect of a single dose of vaccine is not immediate and that not all individuals in the group will be vaccinated in the first week. The pre-vaccination incidence rate ratio (IRR) between the three groups were estimated by Poisson regression based on the weekly incidences of cases (and deaths). Due to over dispersion the confidence intervals for the estimated IRRs were adjusted by a standard error inflation factor. The pre-vaccination IRRs (in relation to the composite control group) were used to prognosticate the post-vaccination incidence of infections (and deaths) in absence of vaccination, by multiplying the pre-vaccination IRRs with the observed post-vaccination incidence in the composite control group, using the confidence intervals of the pre-vaccination IRR to show uncertainty in the resulting estimate. Differences between the prognosticated and observed number of events were used to estimate the number of infections and deaths that may have been prevented up to 2^nd^ May 2021 due to the vaccination programme. Aggregated data on vaccine coverage was grouped by age and residence in LTCF (home care status was not available).

Data was analysed using R version 4.1.0.

### Results

A total 58,174 people (median age 80 years, IQR 71-87 years) were LTCF residents or receiving home care. In the 80+ group there were 62,306 people (median age 83 years, IQR 81-86 years) whilst in the group aged 18-79 there were 1,748,647 people (median age 44 years, IQR 32-58 years). The proportion of men was lower in both the LTCF/home care group and the 80+ group than in the control group 18–79 year-olds (0.40, 0.43, 0.50 respectively) (Table 1).

**Table 1.**
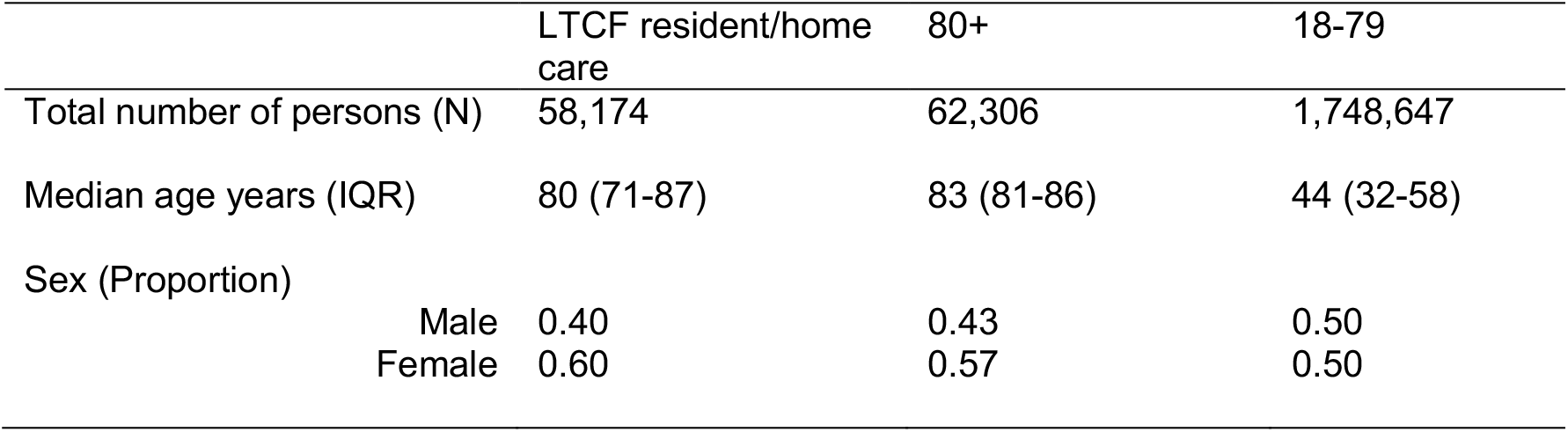
Demographic data for Region Stockholm taken at end of study period 2021-05-02 divided by group

|  | LTCF resident/home care | 80+ | 18-79 |
| --- | --- | --- | --- |
| Total number of persons (N) | 58,174 | 62,306 | 1,748,647 |
| Median age years (IQR) | 80 (71-87) | 83 (81-86) | 44 (32-58) |
| Sex (Proportion) |  |  |  |
| Male | 0.40 | 0.43 | 0.50 |
| Female | 0.60 | 0.57 | 0.50 |

### Infections

The highest number of infections (182,695) were in the control group (18–79 year-olds) (Table 2). More infections were recorded in the LTCF/home care group (6769 total; 5232 pre-vaccination, 1537 post-vaccination) than in the 80+ group (2464 total; 2276 pre-vaccination, 186 post-vaccination). The IRR for SARS-CoV-2 infection in the LTCF/home care group was 1.70 (95%CI 1.54-1.88) pre-vaccination and 0.59 (0.49-0.71) post-vaccination compared to the composite control, higher than for the 80+ group which were 0.38 (0.33-0.44) pre-vaccination and 0.17 (0.09-0.27) post-vaccination (Table 2, Figure 1A). Weekly observed versus expected incidences of infection per 100,000 days at risk are presented in Figure 1A. The estimated number of infections prevented by vaccination was 2873 (2441-3337) for the LTCF/home care group and 239 (178-306) for the 80+ group (Table 2).

**Table 2.** Summary of SARS-CoV2 infections between 2020-08-31 and 2021-05-02 in Region Stockholm

|  | LTCF resident/home care (n=58,174) | 80+ (n= 62,306) | 18-79 (n=1,748,647) |
| --- | --- | --- | --- |
| Total infections during study period (N)* | 6769 | 2462 | 182, 695 |
| Infections after vaccination | 1537 | 186 | N/A |
| Sum expected infections after vaccination | 4410 (3978-4874) | 425 (364-492) | N/A |
| Sum observed infection after vaccination | 1537 | 186 | N/A |
| Estimated prevented infections (95% CI) | 2873 (2441-3337) | 239 (178-306) | N/A |
| Infection incidence rate ratio pre-vaccination (95% CI) | 1.70 (1.54-1.88) | 0.38 (0.33-0.44) | Reference |
| Infection incidence rate ratio post-vaccination (95% CI) | 0.59 (0.49-0.71) | 0.17 (0.09-0.27) | Reference |

\*Summary infections between 2020-08-31 and 2021-05-02

**Figure 1.**
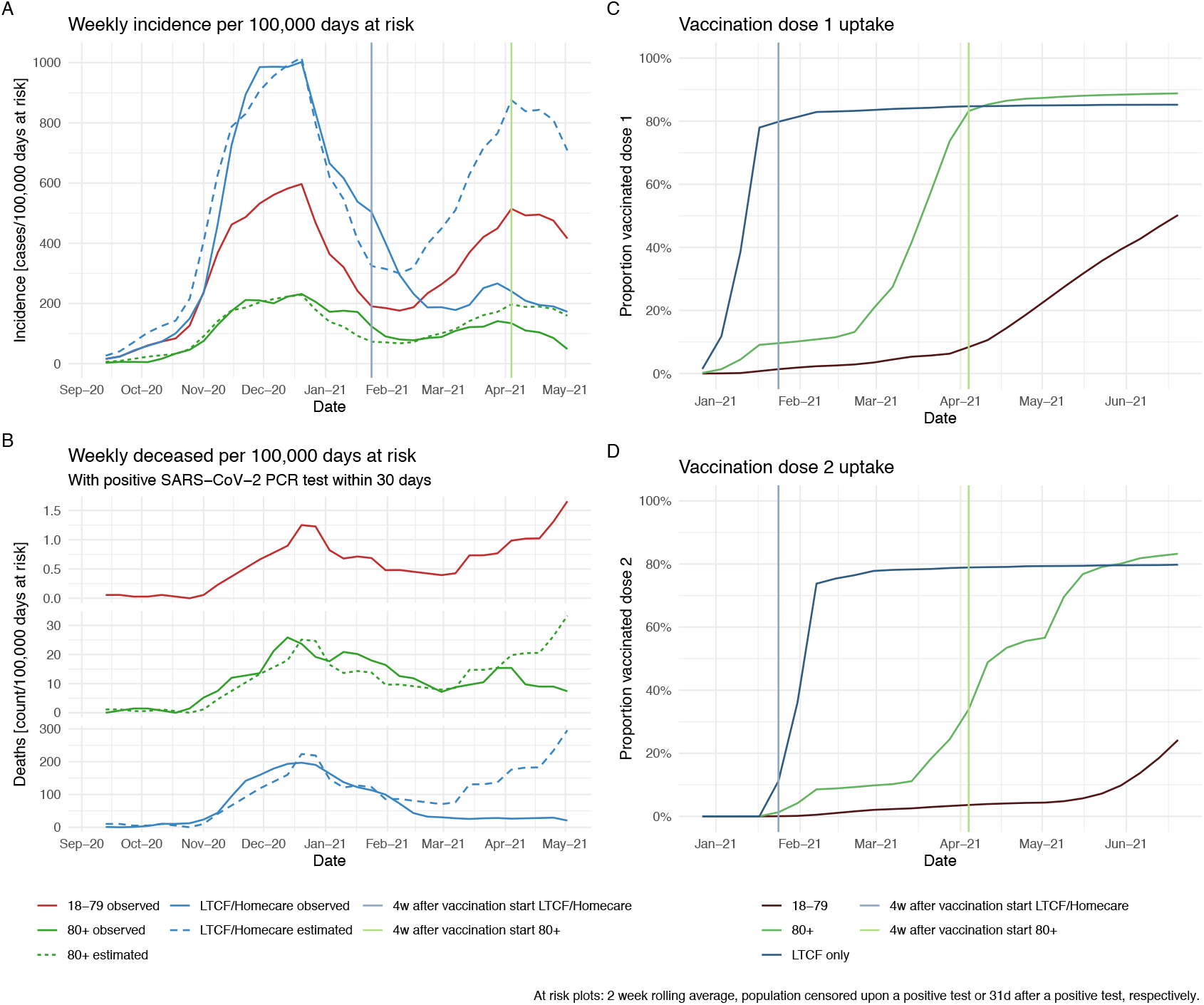
**A**. Weekly incidence of SARS-CoV-2 infection per 100,000 days at risk by group observed and estimated in the absence of vaccination (LTCF/home care, 80+, 18-79) **B**. Weekly deceased per 100,000 days at risk by group observed and estimated in the absence of vaccination (LTCF/home care, 80+, 18-79) **C**. Vaccine coverage with dose 1 by group (LTCF only, 80+, 18-79), **D**. Vaccine coverage with dose 2 by group (LTCF only, 80+, 18-79)

### Mortality

More deaths within 30 days of a positive SARS-CoV2 PCR test were recorded in the LTCF/home care groups than in the 80+ group during the study period (1218 vs 244) (Table 3). In both groups the majority of deaths occurred prior to the start of vaccination; in the LTCF/home care group there were 978 pre-vaccination and 240 post-vaccination whereas in the 80+ group there were 224 pre-vaccination and 20 post-vaccination. 348 deaths were recorded in the 18-79 group. The IRR for death within 30 days of a positive SARS-CoV2 PCR test decreased from 179 (95%CI 146-221) pre-vaccination to 45 (35-59) post-vaccination for the LTCF/home care group and from 20 (16-26) pre-vaccination to 9 (5-18) post-vaccination for the 80+ group, compared to the composite control (Figure 1B). The estimated number of deaths prevented after vaccination was 808 (95%CI 615-1053) for the LTCF/home care group and 46 (32-64) for the 80+ group (Table 3, Figure 1B). Weekly observed versus expected incidence for deaths are presented in Figure 1B.

**Table 3.** Summary of deaths within 30 days of a positive SARS-CoV2 PCR test between 2020-08-31 and 2021-05-02 in Region Stockholm

|  | LTCF resident/homecare<br>(n=58,174) | 80+<br>(n= 62,306) | 18-79<br>(n=1,748,647) |
| --- | --- | --- | --- |
| Total deaths during study period (N) | 1218 | 244 | 348 |
| Deaths after vaccination | 240 | 20 | N/A |
| Death incidence rate ratio pre-vaccination (95% CI) | 179 (146-221) | 20 (16-26) | Reference |
| Death incidence rate ratio post-vaccination (95% CI) | 45 (35-59) | 9 (5-18) | Reference |
| Sum expected deaths after vaccination (95% CI) | 1048 (855-1293) | 66 (52-84) | N/A |
| Sum observed deaths | 240 | 20 | N/A |
| Estimated prevented deaths (95% CI) | 808 (615-1053) | 46 (32-64) | N/A |

### Vaccine coverage and choice of vaccines in Stockholm

Figures 1C and 1D show the vaccine coverage with doses 1 and 2 stratified by LTCF, age 80+ and ages 18-79. In both the LTCF and 80+ group, at least 80% of the group had received a first dose by 4 weeks after the start of vaccination of the group. The vaccine coverage data was based on LTCF and age, since data for those receiving home care was not available by vaccine status.

## Discussion

Our data suggests that vaccination of the LTCF/home care and 80+ groups succeeded in preventing a third wave in these groups of the same magnitude as the control group. Since the start of the vaccination programme there has been no national lock-down or increase in restrictions to visiting friends and relative in LTCF and as such the effect of prevention of infections and deaths is likely to be due to vaccination rather than to new social distancing measures alone[5]. The IRR for infections and deaths was much lower in the 80+ group compared to the LTCF/home care group even prior to vaccination, which is likely to be an effect of the Swedish recommendations, that is, that those over 70 years take extra precautions to avoid social contact with others outside their own household[6]. Those receiving home care who have regular visits by healthcare professionals and those living in LTCF have not been able to isolate to the same extent and have been much more vulnerable to the spread of infection[7].

A combination of EU approved vaccines has been used since the start of the programme. Use of the Pfizer-BioNTech mRNA vaccine started on 27^th^ December 2020 followed by the Moderna mRNA vaccine after it received EMA approval on 6^th^ January 2021[8]. Astra-Zeneca’s COVID-19 vaccine received EMA approval on 29^th^ January 2021[8] although vaccination of those over 65 years with Astra-Zeneca’s vaccine only started on 4^th^ March due to initial concerns about effectiveness in older adults[9]. Vaccination with Astra-Zeneca’s vaccine was paused between 16^th^ and 25^th^ March whilst reports of links with a rare blood clotting disorder were investigated[3]. Since 25^th^ March, vaccination with Astra-Zenecas vaccine has been resumed amongst those over 65[3, 10, 11]. It is likely therefore that the majority of those vaccinated in the LTCF/home care and 80+ groups received mRNA vaccines.

The effect observed is a combination of protection from a single dose and two doses. High vaccine coverage by 4 weeks post-vaccination was achieved in both of the first groups targeted for vaccination. Timing of the second dose of vaccinations has varied in Stockholm based on which vaccine is given and time point during the vaccination programme. Currently the mRNA vaccines are given with a 3-7 week dosing interval and Astra-Zeneca’s vaccine with a 6-12 week interval[12].

This study was limited to programmatic effect in that we were unable to link individual level vaccine data as has recently been done in Israel[13] however little has yet been published on the impact of programmes using heterologous vaccines such as this one which is one of the studies strengths. A recent internal report from Public Health England estimates 10,400 deaths have been prevented in England in the first 4 months of their COVID-19 vaccination programme using non-individual level data[14]. The advantage of before and after studies is that they take into account both the direct and indirect effects of vaccination.

An additional limitation in our study is that vaccine coverage data was based on LTCF and age, since data for those receiving home care was not available whereas incidence of infections and deaths were calculated for the combined group (LTCF/home care). Some of the reduction observed in the LTCF/home care group is likely due to indirect effects of vaccination of healthcare workers. Reductions in observed vs. expected infections in the 80+ group started before vaccination indicating that the assumptions about symmetrical infection incidence ratio during the second and third wave might not be fully accurate. The overall trend, however, following initiation of vaccination is highly encouraging.

In conclusion, the programmatic early effects of vaccination in two of the most vulnerable groups are both reduced SARS-CoV-2 infections and deaths.

## Data Availability

Data was extracted from the VAL database in Region Stockholm.

## Authorship

All authors have seen and approved the manuscript and contributed significantly to the work. PN devised the study, CI wrote the manuscript and contributed to the analysis, DS performed the main analysis, FG assisted with statistical analyses and MPH analysed and provided the vaccination data.

## References

[1] Folkhälsomydigheten, Nationell plan för vaccination mot covid-19 (delrapportering 3) 2021. https://www.folkhalsomyndigheten.se/contentassets/43a1e203f7344a399367b816e2c7144c/nationell-plan-vaccination-covid-19-delrapport-3.pdf. (Accessed 4th July 2021).

[2] R. Stockholm, 17 juni: Dagsläge covid-19, 2021. https://www.sll.se/verksamhet/halsa-och-vard/nyheter-lagesrapporter-covid-19/2021/06/17-juni-dagslage-covid-19/. (Accessed 19th June 2021).

[3] Folkhälsomydigheten, Information om vaccination med Astra Zenecas vaccin till personer som är 65 år och äldre, 2021. https://www.folkhalsomyndigheten.se/smittskydd-beredskap/utbrott/aktuella-utbrott/covid-19/vaccination-mot-covid-19/om-vaccinerna-mot-covid-19/information-om-fortsatt-vaccination-av-astra-zenecas-vaccin-till-personer-som-ar-65-ar-och-aldre/. (Accessed 4th June 2021).

[4] T. Forslund, A.C. Carlsson, G. Ljunggren, J. Arnlov, C. Wachtler, Fam Pract 38(2) (2021) 132–140.

[5] Folkhälsomydigheten, Tillfälliga lokala besöksförbud på äldreboenden upphör, 2021. https://www.folkhalsomyndigheten.se/smittskydd-beredskap/utbrott/aktuella-utbrott/covid-19/information-till-varden/personal-inom-aldreomsorg/lokala-besoksforbud-pa-aldreboenden/. (Accessed 1st July 2021).

[6] Folkhälsomydigheten, Folkhälsomyndighetens föreskrifter och allmänna råd om allas ansvar att förhindra smitta av covid-19 m.m., 2020. https://www.folkhalsomyndigheten.se/contentassets/a1350246356042fb9ff3c515129e8baf/hslf-fs-2020-12-allmanna-rad-om-allas-ansvar-covid-19-tf.pdf. (Accessed 4th July 2021).

[7] S. Baral, R. Chandler, R.G. Prieto, S. Gupta, S. Mishra, M. Kulldorff, Ann Epidemiol 54 (2021) 21–26.

[8] Läkemedelsverket, Coronavaccin, vaccin mot coronaviruset (covid-19), 2021. https://www.lakemedelsverket.se/sv/coronavirus/coronavaccin. (Accessed 4th July 2021).

[9] Folkhälsomydigheten, Samtliga godkända vacciner mot covid-19 skyddar mot sjukdom, 2021. https://www.folkhalsomyndigheten.se/nyheter-och-press/nyhetsarkiv/2021/februari/samtliga-godkanda-vacciner-mot-covid-19-skyddar-mot-sjukdom/. (Accessed 4th July 2021).

[10] Folkhälsomydigheten, Om vaccinerna mot covid-19, 2021. https://www.folkhalsomyndigheten.se/smittskydd-beredskap/utbrott/aktuella-utbrott/covid-19/vaccination-mot-covid-19/om-vaccinerna-mot-covid-19/om-vaccinerna/. (Accessed 4th June 2021).

[11] Folkhälsomydigheten, Rekommendation om åldersgräns på 65 år för AstraZenecas vaccin kvarstår, 2021. https://www.folkhalsomyndigheten.se/nyheter-och-press/nyhetsarkiv/2021/april/rekommendation-om-aldersgrans-pa-65-ar-for-astrazenecas-vaccin-kvarstar/. (Accessed 4th June 2021).

[12] Folkhälsomydigheten, Om vaccinerna mot covid-19, 2021. https://www.folkhalsomyndigheten.se/smittskydd-beredskap/utbrott/aktuella-utbrott/covid-19/vaccination-mot-covid-19/om-vaccinerna-mot-covid-19/om-vaccinerna/. (Accessed 4th July 2021).

[13] E.J. Haas, F.J. Angulo, J.M. McLaughlin, E. Anis, S.R. Singer, F. Khan, N. Brooks, M. Smaja, G. Mircus, K. Pan, J. Southern, D.L. Swerdlow, L. Jodar, Y. Levy, S. Alroy-Preis, Lancet 397(10287) (2021) 1819–1829.

[14] J.S. Nick Andrews, Sharif Ismael, Laura Coughlan, Hester Allen, Mary Ramsay, Jamie Lopez Bernal, Impact of COVID-19 vaccines on mortality in England: December 2020 to March 2021, 2021. https://assets.publishing.service.gov.uk/government/uploads/system/uploads/attachment_data/file/977249/PHE_COVID-19_vaccine_impact_on_mortality_March.pdf. (Accessed 4th July 2021).

